# Large-scale proteomics in early pregnancy and timing of onset of hypertensive disorders of pregnancy

**DOI:** 10.64898/2026.06.09.26355317

**Authors:** Alisse Hauspurg, Xiaoning Huang, Philip Greenland, Victoria Pemberton, Noel Bairey Merz, George Saade, Lynn M. Yee, Lisa D. Levine, Angela Ranzini, David Haas, Matthew Hoffman, Emily Lau, Sadiya S. Khan, Brian Kleiboeker, Uma Reddy, Janet Catov, William Grobman

## Abstract

**Background:** Hypertensive disorders of pregnancy (HDP) may first be diagnosed antepartum, during labor, or postpartum. We utilized untargeted large-scale proteomics to identify pathways associated with HDP based on timing of onset.

**Methods:** We performed a nested case-control study comparing differential protein expression, from the SomaScan 7K platform, based on timing of onset of HDP (categorized as antepartum, intrapartum, postpartum) versus controls (referent) using first-trimester plasma samples from the NuMoM2b-Heart Health Study, a multi-site prospective cohort that followed nulliparous individuals from the first trimester through postpartum. Associations of individual proteins with timing of onset of HDP, adjusted for co-variates, were assessed using logistic regression q value-based false discovery rates and pathway enrichment and differential expression analysis were conducted.

**Results:** Of 1628 individuals included, 678 had a HDP, of which 67% initially manifested antepartum (AP), 29% intrapartum (IP), and 3% postpartum (PP). After adjusting for co-variates, compared to controls, 698 proteins, 39 proteins, and 144 proteins were differentially expressed in those with HDP according to AP, IP, PP onset, respectively. There was little overlap in individual protein expression based on timing of HDP diagnosis, with pathway enrichment analysis and graphical summary analysis suggesting distinct processes. Specifically, there was downregulation of angiogenic proteins in AP HDP, downregulation of immune-related proteins in IP HDP, and upregulation of complement activation promoting fibrotic and inflammatory changes leading to cardiac dysfunction in PP HDP.

**Conclusion:** There are differences in first-trimester protein expression based on whether HDP first manifests AP, IP or PP. This raises the possibility that there may be distinct mechanistic phenotypes that could uniquely inform diagnostic and therapeutic targets for HDP.

## INTRODUCTION

Hypertensive disorders of pregnancy (HDP), including preeclampsia and gestational hypertension, can first manifest at different times during pregnancy: antepartum (during pregnancy but before labor), intrapartum (during labor), or postpartum (in the first six weeks after delivery).^1^ Over 40% of HDP is diagnosed in the intrapartum and postpartum periods, yet HDP during these times has largely been understudied in epidemiologic and translational investigations.^2^ Indeed, sometimes it is not even labelled as HDP given that new-onset elevated blood pressure during labor or postpartum is frequently attributed to pain or catecholamine release.^3^ We previously demonstrated that new-onset HDP is associated with an increased risk of incident hypertension in the 2-7 years following delivery regardless of whether it was diagnosed antepartum, intrapartum, or postpartum.^2,4^ However, the magnitude of risk of incident hypertension differs by timing of HDP onset, suggesting the possibility that there may be distinct underlying pathophysiologic pathways.^2^

The concept that HDP may comprise distinct pathophysiologic entities with differing mechanistic underpinnings is relatively well established for HDP classified by gestational age of onset, with many studies distinguishing between early-onset (<34 weeks of gestation) and late-onset (>34 weeks of gestation) disease.^5,6^ These subtypes, moreover, have been shown to exhibit distinct proteomic signatures in early pregnancy.^7,8^ However, whether HDP arising during labor (intrapartum) or after delivery (postpartum) represent pathophysiologically distinct entities from that which arises antepartum remains unclear.

We hypothesized that timing of onset of HDP in a first pregnancy is reflective of different mechanisms that can be identified by their proteomic signatures in early pregnancy. In the current study, our goal was to determine whether differential proteomic expression patterns in first trimester serum were associated with timing of HDP onset in a diverse cohort of nulliparous individuals.

## MATERIALS AND METHODS

This is a secondary analysis of the Nulliparous Pregnancy Outcomes Study: Monitoring Mothers-to-Be (nuMoM2b), a National Institutes of Health–funded study that recruited nulliparous individuals with singleton pregnancies from 8 hospital systems in the US from 2010 to 2013. Details of the methods have been previously described.^9^ Briefly, participants were eligible for enrollment if they had a singleton gestation with cardiac activity, were between 6 weeks 0 days and 13 weeks 6 days, and had no prior pregnancy lasting 20 weeks or more. Participants had study visits once during each trimester and again at delivery. Blood samples used for this analysis were collected during the first-trimester study visit.

### Cases and Controls

This study utilizes a nested case-control design. Individuals with stored plasma specimens from the first-trimester visit who delivered at or after 20 weeks gestation with an HDP (defined as gestational hypertension or preeclampsia) were defined as cases. Cases were further subdivided by whether HDP onset was antepartum, intrapartum, or postpartum (indicated as antepartum-onset, intrapartum-onset or postpartum-onset). Controls were randomly selected from participants who delivered at or after 37 weeks without any HDP, preterm birth, or small for gestational age. Individuals with pre-pregnancy chronic hypertension were excluded from this analysis. To allow for future longitudinal analysis, participant selection was limited to those who attended a follow-up study visit 2 to 7 years after delivery.

### Proteomics Methods

The SomaScan assay (SomaLogic) and its performance characteristics have been reported previously.^10^ This assay uses DNA-based binding reagents (modified aptamers) to quantify the availability of binding epitopes on plasma proteins with high specificity and limits of detection largely comparable to antibody-based assays.^10–12^ The assay starts as a mixture of thousands of fluorophore-labeled modified aptamer reagents immobilized on streptavidin-coated beads and incubated with 55μL of EDTA plasma. Samples are run at three different dilutions to expand dynamic range to approximately 10 logarithms. After a series of washing steps and use of a polyanionic competitor to negate nonspecific binding, and a second capture step, modified aptamer reagents are hybridized to complementary sequences on a DNA microarray chip and quantified by fluorescence, which is related to the relative availability of each protein in the original sample. We used version 4.1 of the SomaScan, which includes 7596 aptamers. As in prior studies, we excluded aptamers paired to nonhuman proteins and aptamers that were incompletely characterized.^13–15^ This left 7335 aptamers and 6481 unique proteins (some proteins are measured by 2 or more aptamers). Median intraassay coefficients of variation for SomaScan of plasma samples are 5% or less.^16,17^ SomaLogic performs an internal quality control analysis (https://dam.somalogic.com/m/34de2ede028f756b/original/D00006601_Rev-1_2023-12_Flagged-Sample-Tech-Note.pdf), and samples were excluded that did not pass quality control.

### Outcomes and covariates

Pregnancy outcomes, including timing of onset of HDP, were prospectively collected and validated using medical record abstraction; methods and definitions have been reported previously.^2,9^ The primary outcome for this analysis was presence and timing of onset of HDP during the nuMoM2b pregnancy: no HDP, antepartum-onset HDP, intrapartum-onset HDP or postpartum-onset HDP. Covariates used in the multivariate model, chosen *a priori*, were age, self-reported race and ethnicity (considered a social construct), body mass index (BMI), diabetes, health insurance, and fetal sex.

### Statistical Analysis

All statistical analyses and data visualization were performed in Stata version 19. Demographic and baseline clinical characteristics are described using means and standard deviations for continuous variables or counts and percentages for categorical variables. Differential expression analysis across timing of HDP groups compared to controls was conducted. Associations of individual proteins with HDP by timing of onset, both unadjusted and adjusted for covariates, were assessed using logistic regression and attendant volcano plots and q value-based false discovery rates (FDR). For single-protein analyses, we accounted for multiple testing across all proteins using Benjamini–Hochberg false discovery rate correction. Proteins with FDR-adjusted q values < 0.05 were considered statistically significant. For multiple comparisons (e.g., differential expression analysis), q values (FDR-adjusted p-values) < 0.05 were used to determine significance.

Pathway enrichment analysis was used to group differentially expressed proteins according to molecular functions, biologic processes, and protein class. Ingenuity Pathway Analysis (IPA; QIAGEN Inc) was performed to discover biological processes regulated by the proteins of interest, and to identify canonical pathways significantly upregulated or downregulated compared to controls across each of the types of HDP. Pathways of interest were identified with an absolute value z score > 2. Finally, IPA was used to generate graphical summaries of the top enriched and predicted terms in the form of a causal network.

## RESULTS

Proteomic assay results were analyzable for 1628 participants. Among those, 950 had normotensive pregnancies and 678 had a pregnancy with HDP, with 456 (67%) having antepartum-onset HDP, 200 (29%) having intrapartum-onset HDP, and 22 (3%) having postpartum-onset HDP. Compared to those with normotensive pregnancies, those with HDP had a higher early pregnancy BMI and higher systolic and diastolic blood pressure (BP) in early pregnancy. Demographic and obstetric characteristics are summarized in Table 1.

**Table 1.** Demographic and pregnancy characteristics of participants by timing of onset of HDP.

|  | **Normotensive pregnancy** | **Antepartum HDP** | **Intrapartum HDP** | **Postpartum HDP** | **p-value** |
| --- | --- | --- | --- | --- | --- |
|  | **(n= 950)** | **(n= 456)** | **(n=200)** | **(n=22)** |  |
| Age at delivery (years) | 27.2 (5.4) | 26.8 (5.6) | 27.0 (5.8) | 27.8 (6.1) | 0.62 |
| Early pregnancy BMI (kg/m<sup>2</sup>) | 24.4 (22.0, 28.8) | 28.2 (23.8, 33.6) | 25.9 (22.7, 31.0) | 26.0 (23.7, 30.1) | <0.001 |
| Race |  |  |  |  |  |
| Non-Hispanic White | 597 (62.8%) | 301 (66.0%) | 107 (53.5%) | 12 (54.5%) | <0.001 |
| Non-Hispanic Black | 117 (12.3%) | 73 (16.0%) | 46 (23.0%) | 6 (27.3%) |  |
| Hispanic | 168 (17.7%) | 46 (10.1%) | 27 (13.5%) | 3 (13.6%) |  |
| Asian | 27 (2.8%) | 10 (2.2%) | 5 (2.5%) | 0 (0.0%) |  |
| Other | 41 (4.3%) | 26 (5.7%) | 15 (7.5%) | 1 (4.5%) |  |
| Early pregnancy SBP (mmHg) | 108.1 (10.1) | 114.3 (10.5) | 109.5 (10.9) | 112.9 (12.6) | <0.001 |
| Early pregnancy DBP (mmHg) | 66.8 (7.9) | 71.0 (8.7) | 68.1 (8.7) | 67.0 (8.4) | <0.001 |
| Gestational diabetes | 86 (9.1%) | 26 (5.7%) | 18 (9.0%) | 1 (4.6%) | <0.001 |
| Gestational age at delivery (weeks) | 39.3 (1.1) | 37.8 (2.8) | 38.5 (3.0) | 38.3 (2.3) | <0.001 |

After adjusting for covariates, compared with controls, those with HDP had 698 proteins, 39 proteins, and 144 proteins differentially expressed based on whether HDP was AP, IP, PP onset, respectively with FDR-adjusted q value <0.05 (Figure 2). Supplemental Tables 1-3 provide related summaries for the 30 most statistically significant proteins by group. There was little overlap in individual protein expression based on timing of HDP onset compared to controls (Figure 3). In total, there were 4 proteins identified that overlapped in those who developed antepartum versus intrapartum onset and 14 identified that overlapped in those who subsequently developed antepartum versus intrapartum onset. There were no proteins identified that overlapped in those who subsequently developed HDP intrapartum versus postpartum onset. Supplemental Tables 4 and 5 provide summaries for overlapping proteins between groups.

**Figure 2.**
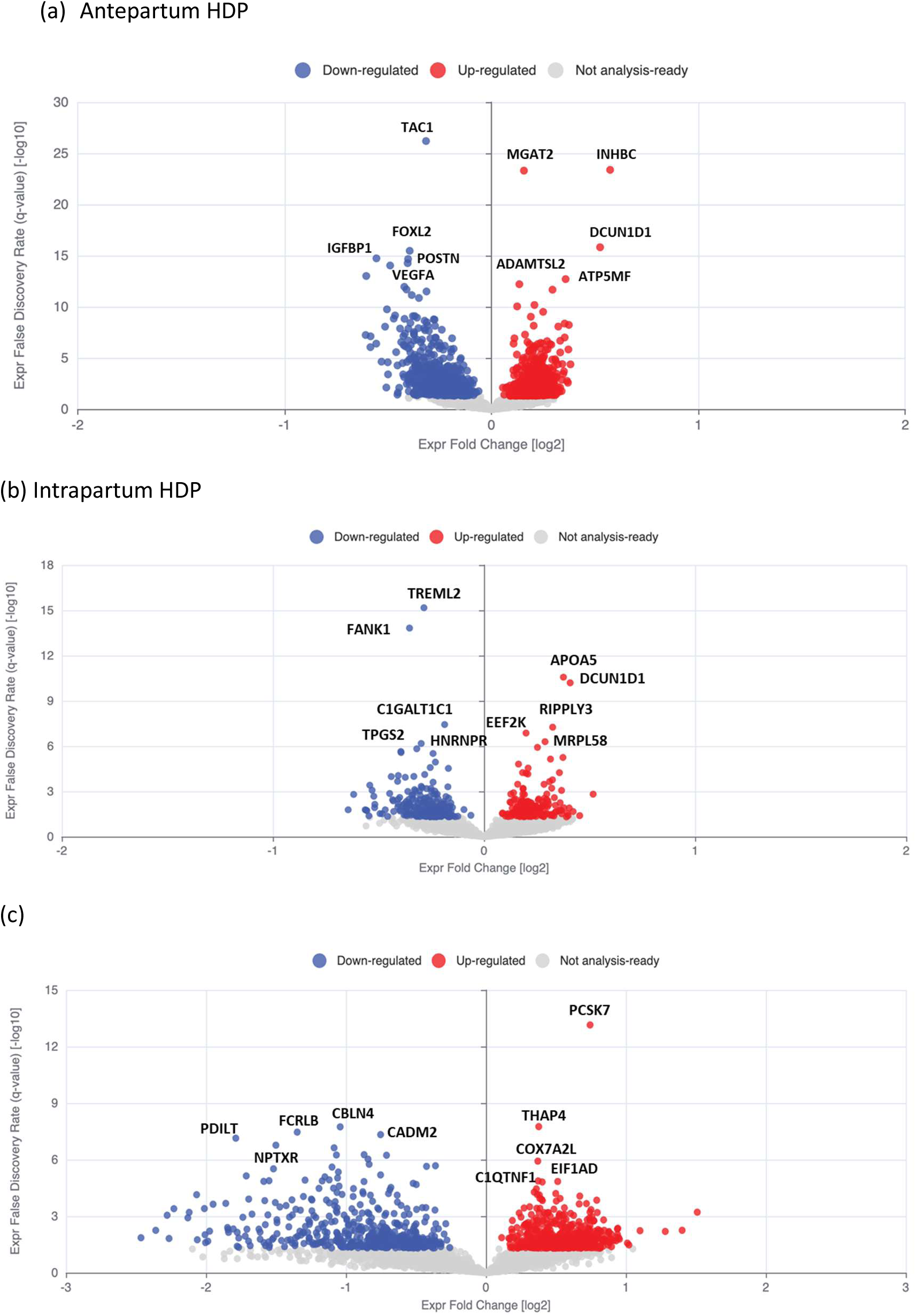
Volcano plot of proteins associated with HDP with varying timing of onset (a) antepartum, (b) intrapartum and (c) postpartum compared to controls using first trimester plasma samples, after co-variate adjustment.

**Figure 3.**
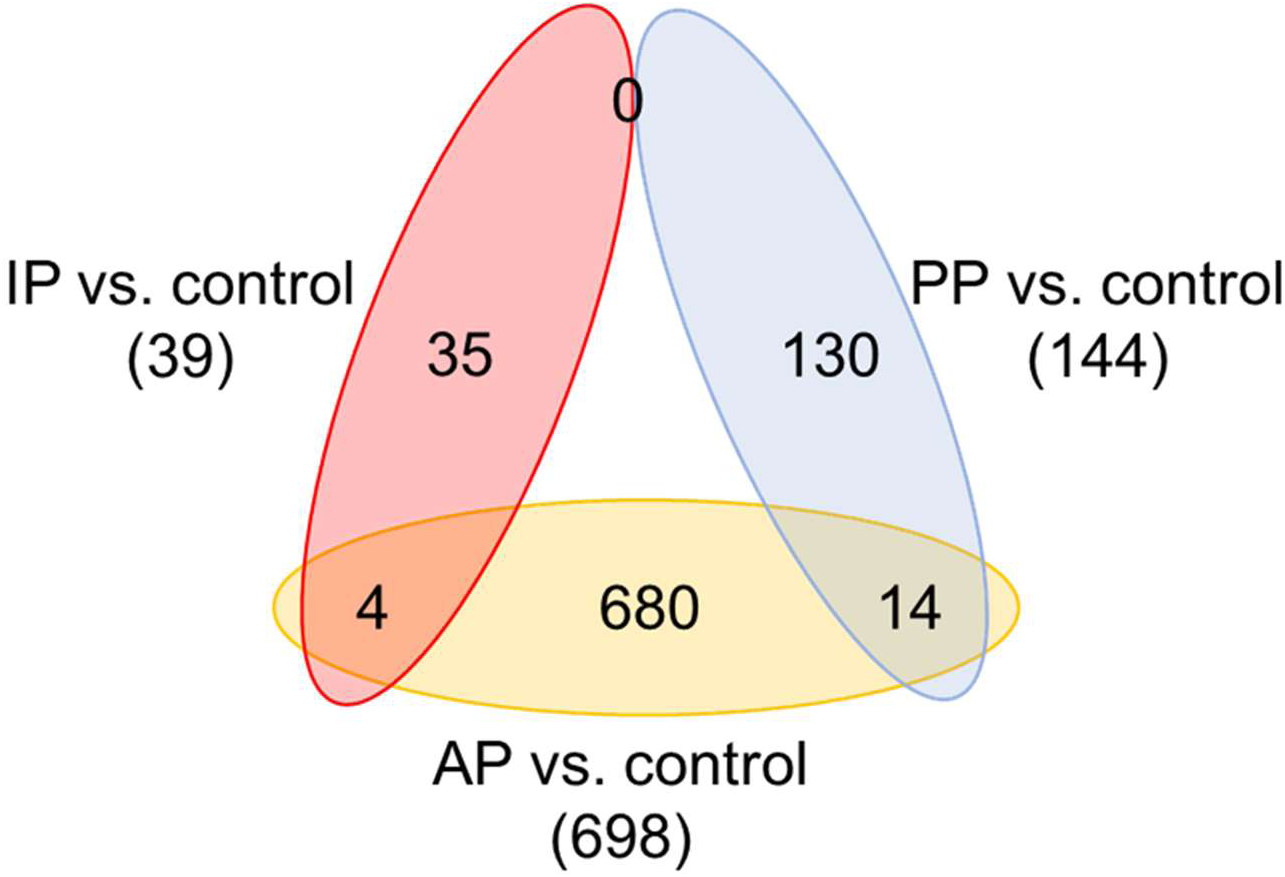
Venn diagram depicting the overlap of differentially expressed proteins between each timing of onset group (antepartum [AP], intrapartum [IP], and postpartum [PP]) and controls.

The canonical pathways, identified by IPA, that were significantly upregulated or downregulated compared to controls by type of HDP are presented in Figure 4. For antepartum HDP, the most significantly upregulated pathway was “Signaling by VEGF”. Upregulation of this pathway leads to downregulation of VEGFA, fms-related receptor tyrosine kinase 1 (*FLT1*), neuropilin 2 and placental growth factor (*PlGF*). The most significantly downregulated pathway involved the “Role of Osteoblasts in Rheumatoid Arthritis Signaling Pathway”, with the most downregulated proteins in the pathway including vascular endothelial growth factor A (*VEGFA*) and matrix metallopeotidase 12 (*MMP12*). In contrast, for intrapartum HDP, the most significantly upregulated pathway was “SPINK1 Pancreatic Cancer Pathway*”,* which is often activated in the setting of inflammation, and the most significantly downregulated pathway was “Neutrophil degranulation”. Finally, for postpartum HDP, the most significantly upregulated pathway was “Regulation of the Epithelial Mesenchymal Transition by Growth Factors*,”* which involves processes such as wound healing, tissue-regeneration and fibrosis. The most significantly downregulated pathway associated with postpartum HDP was “Airway Pathology in Chronic Obstructive Pulmonary Disease”, which involves activation of transforming growth factor beta (*TGF-β*) and fibroblast activation leading to extracellular matrix accumulation.

**Figure 4.**
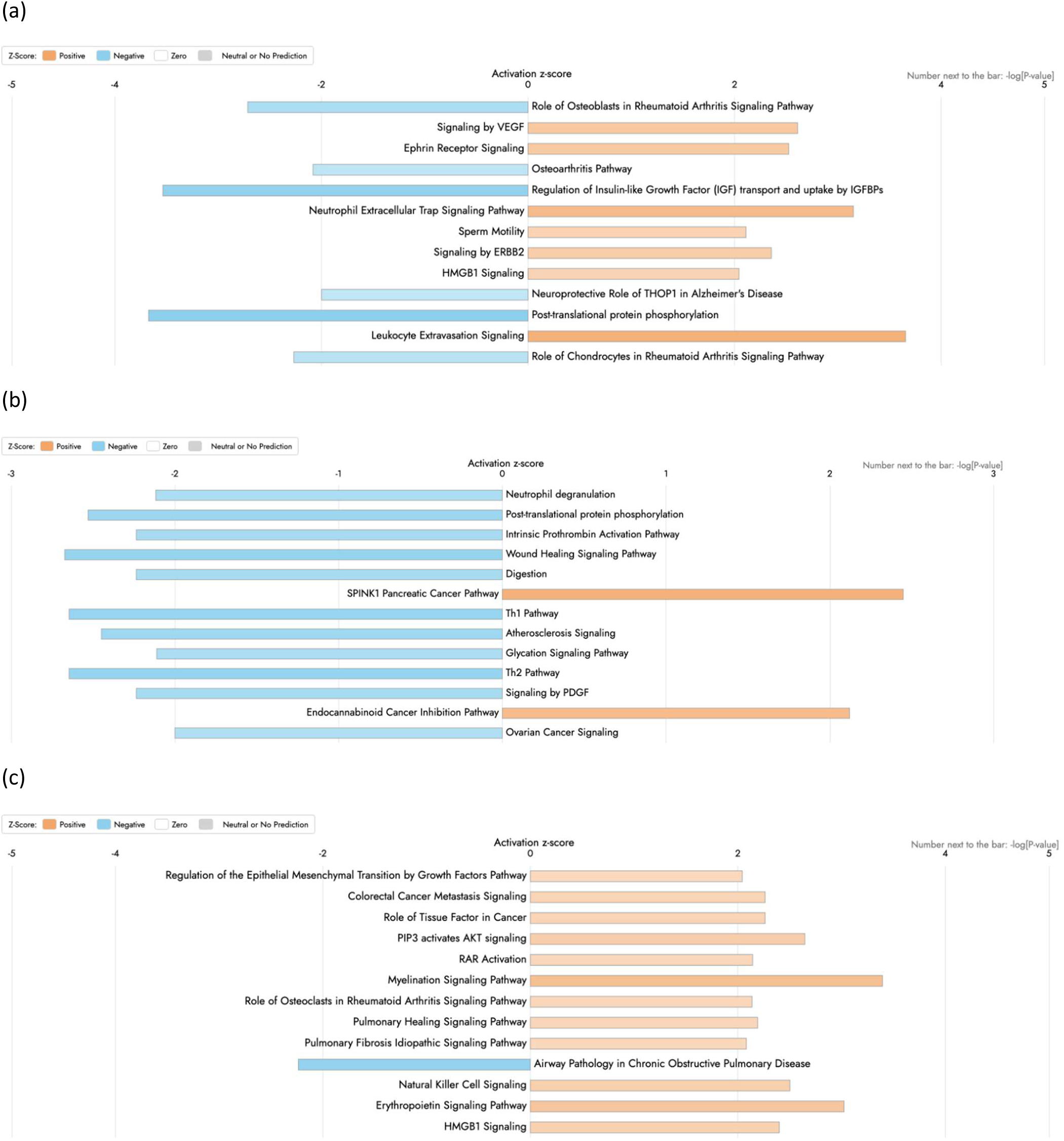
Ingenuity pathway analysis (IPA) of proteins dysregulated in HDP with varying timing of onset relative to controls (absolute value z score >2). Canonical pathways dysregulated in HDP with timing of onset (a) antepartum (b) intrapartum and (c) postpartum with P < 0.05 are sorted by p-value. The color indicates the direction of expression change: red indicates upregulation and blue indicates downregulation relative to control. Color intensity reflects the magnitude of fold change.

Graphical summary analyses are shown in Figure 5. The key themes that emerge for antepartum HDP include downregulation of vasculogenesis and upregulation of fibroblast transformation and fatty acid metabolism. In contrast, intrapartum HDP involves downregulation of many immune-related processes and regulation of insulin-like growth factor. Postpartum HDP involves several upregulated pathways that converge on “Dysfunction of heart”, with complement activation (*C3*) that promotes fibrotic and inflammatory changes leading to cardiac dysfunction.

**Figure 5.**
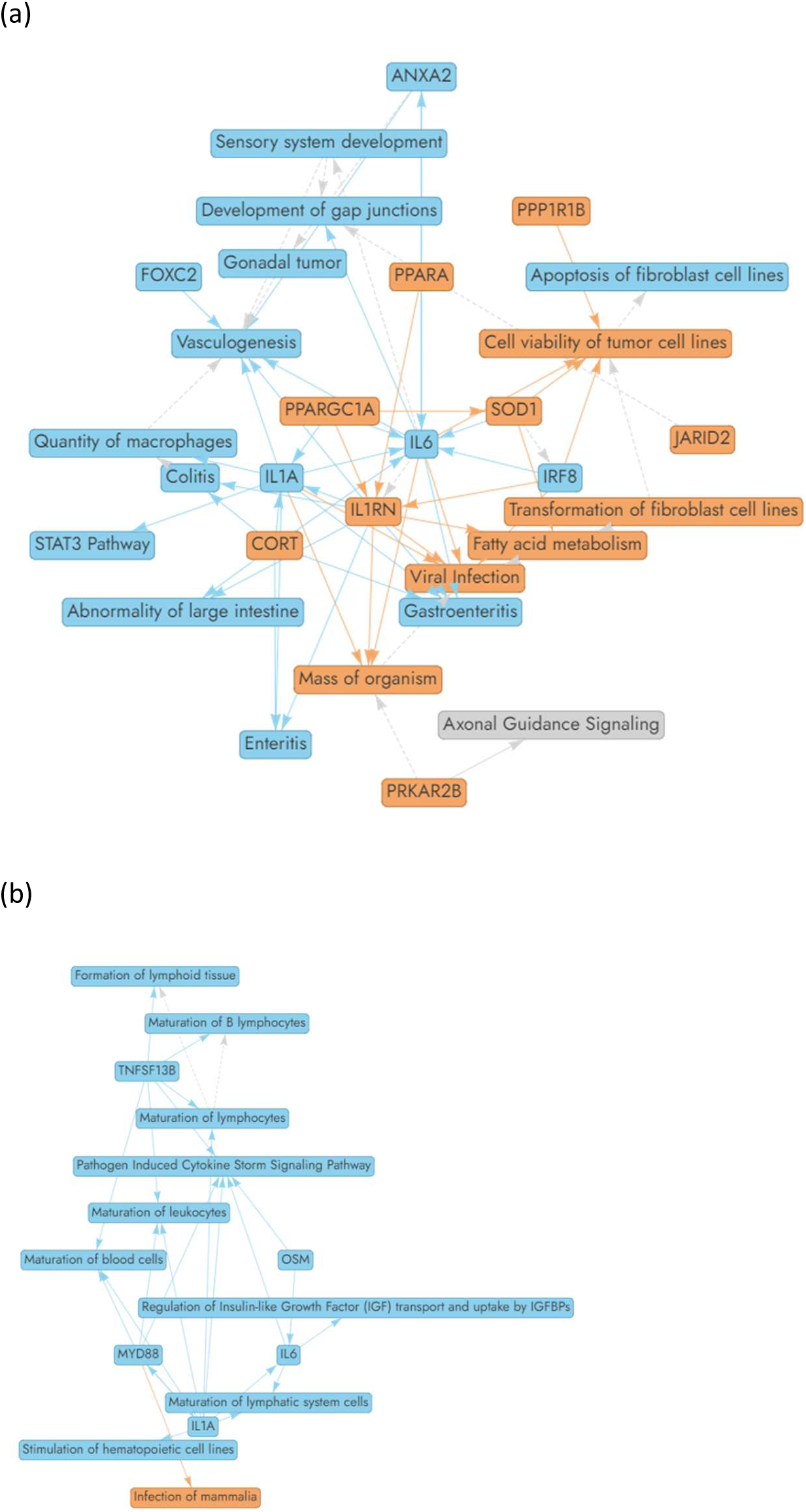
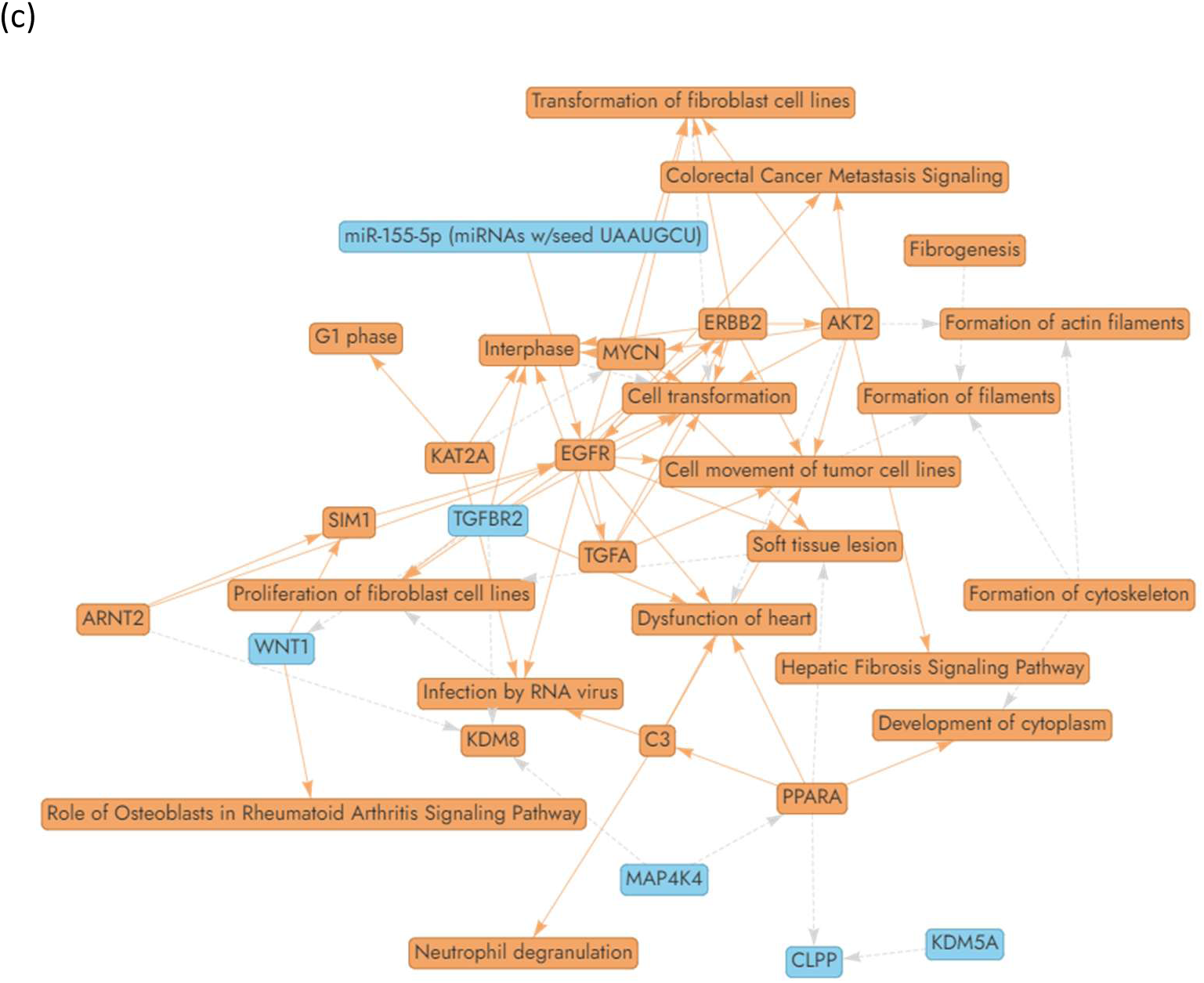
Ingenuity pathway analysis (IPA) of proteins dysregulated in HDP with varying timing of onset relative to controls. Graphical summary analysis depicting HDP with timing of onset (a) antepartum (b) intrapartum and (c) postpartum. The color indicates the direction of expression change: red indicates upregulation and blue indicates downregulation relative to control.

## DISCUSSION

In this large-scale proteomic analysis of first-trimester samples collected from nulliparous individuals enrolled in a multi-site prospective cohort study, we identified distinct proteomic signatures associated with subsequent antepartum, intrapartum, and postpartum-onset of HDP. The minimal overlap in differentially expressed proteins across the groups supports the hypothesis that timing of onset of HDP may reflect distinct pathophysiologic phenotypes that are potentially distinguishable as early as the first trimester of pregnancy. Furthermore, pathway enrichment analyses demonstrated fundamentally different biological processes that may underlie each HDP subtype: antepartum HDP was characterized by dysregulated VEGF signaling, intrapartum HDP by inflammatory and immune-related pathways, and postpartum HDP by complement-mediated cardiac dysfunction pathways.

The finding that antepartum HDP is most prominently characterized by VEGF signaling dysregulation, including downregulation of *VEGFA, FLT1*, neuropilin 2, and *PlGF* is consistent with our current understanding of the pathogenesis of preeclampsia, in which abnormal trophoblast invasion leads to impaired spiral artery remodeling, placental ischemia, and subsequent release of anti-angiogenic factors.^18–20^ One such factor is soluble fms-like tyrosine kinase-1 (*sFLT1*), which neutralizes *VEGF* and *PlGF* resulting in endothelial dysfunction.^19,21^ The detection of angiogenic imbalance in first-trimester samples in this study extends prior work showing that circulating angiogenic markers are altered weeks before clinical disease onset and aligns with the concept that antepartum HDP, particularly early-onset disease, represents a primarily placentally-driven phenotype.^22,23^ The concurrent downregulation of *MMP12* in the osteoblast-related pathway further suggests early disruption of extracellular matrix remodeling, which is critical for normal placental vascular development.^24^ Notably, the antepartum group yielded the largest number of differentially expressed proteins (n=698), consistent with the expectation that placental-origin disease would produce the most robust early proteomic signal, as the abnormal spiral artery remodeling typically occurs early in gestation. While consistent with prior studies, replication of these findings in a large, multi-site cohort provides important external validation and supports previously published work about the hypothesized mechanisms.^18,20,21,23,25,26^

In contrast, the proteomic signature of intrapartum HDP was markedly different, with the most significantly upregulated pathway being the “SPINK1 Pancreatic Cancer Pathway”. *SPINK1* is a serine protease inhibitor that functions as an acute-phase reactant and is upregulated in inflammatory states through Nuclear Factor kappa-light-chain-enhancer of activated B cells (*NF-κB)* and Interleukin-6 / Signal Transducer and Activator of Transcription 3 (*IL-6/STAT3)* signaling.^27–29^ First-trimester activation of *SPINK1* in those who subsequently develop intrapartum HDP suggests that this phenotype may be dependent on an exaggerated inflammatory response rather than the angiogenic imbalance characteristic of antepartum disease. These findings challenge the clinical assumption that new-onset intrapartum hypertension is primarily related to pain or a stress response and instead suggest, at least for some individuals, a distinct inflammatory phenotype identifiable by antecedent molecular changes. The relatively small number of differentially expressed proteins in the intrapartum group (n=39) may reflect a more subtle or later in gestation pathophysiologic process that is less manifest in the first trimester or may indicate that intrapartum HDP is more etiologically heterogeneous.

Postpartum HDP demonstrated yet another distinct pathway profile and phenotype, with upregulation of “Regulation of Epithelial Mesenchymal Transition by Growth Factors” and downregulation of pathways involving *TGF-β* and fibroblast activation. Epithelial-mesenchymal transition (EMT) and its endothelial counterpart (EndMT) are processes by which cells acquire mesenchymal characteristics, contributing to fibrosis, wound healing, and vascular remodeling.^30,31^ *TGF-β* is a key regulator of EndMT and has been implicated in cardiac fibrosis and atherosclerosis.^30,32^ The graphical summary analysis further revealed that postpartum HDP involves upstream regulators converging on the “Dysfunction of heart” pathway, with complement activation (C3) promoting fibrotic and inflammatory changes leading to cardiac dysfunction. Complement C3 activation previously has been shown to regulate cardiac remodeling and is dysregulated in heart failure, and the C3-complement factor D-C3a receptor axis has been identified as a critical mediator of right ventricular failure.^33,34^ These findings suggest that postpartum HDP may represent a phenotype more closely aligned with subclinical cardiac dysfunction rather than with traditional placental disease, which is supported by the observation that individuals with HDP in the postpartum period often have significant volume overload and heart failure symptoms, with a clinical presentation that resembles heart failure with preserved ejection fraction (HFpEF).^35,36^ Recent proteomic work, which aligns with our results, has identified placental senescence and the senescence-associated secretory phenotype (SASP), including activin A and *TGF- β* family members, as shared pathways in both peripartum cardiomyopathy and preeclampsia.^37^

Our findings build upon a prior analysis from this cohort, which evaluated first-trimester proteomic analysis associated with HDP overall. Our analysis, which explored how proteomic signatures associated with HDP vary by timing of onset, suggests the potential importance of stratifying by the timing of disease onset.^6^ This finding may have important implications for future exploration of biomarkers, as prediction models trained on HDP overall, or antepartum HDP alone, may perform poorly for intrapartum and postpartum subtypes. The different mechanistic pathways associated with each subtype suggest that prevention strategies may need to be tailored to HDP subtype. For example, interventions targeting angiogenic imbalance (e.g., aspirin, which has been shown to reduce preterm preeclampsia)^38^ may be most effective for prevention of antepartum-onset disease, while anti-inflammatory or immunomodulatory approaches might be more appropriate for prevention or treatment of intrapartum-onset HDP, and cardio-protective strategies may be most appropriate for prevention of postpartum-onset HDP. These insights may also extend to postpartum interventions to improve long-term maternal health after HDP.

There are several strengths of our study. Compared to previous proteomic studies, our cohort is larger, more diverse, and used an aptamer-based proteomics method that included over 6400 proteins. Outcome assessment was standardized through the study protocol with chart review and adjudication by trained abstracters based on signs and symptoms and not only whether HDP was diagnosed at the time of clinical care or captured in administrative codes. This study also has several limitations. Our cohort is comprised of nulliparous individuals; thus, our findings may not be applicable to multiparous individuals. Additionally, while we adjusted for key co-variates associated with HDP, residual confounding cannot be excluded. The postpartum-onset group was relatively small (n=22), which limits statistical power and pathway-level inferences for this subgroup. Additionally, as with all studies of timing of hypertension onset, the timing of diagnosis may not truly reflect timing of onset, such as in cases in which individuals with intrapartum hypertension had unrecognized hypertension in the period prior to labor. Further, this study lacks a validation cohort, and as such, we consider our results to be hypothesis-generating, which require validation in additional, larger cohorts. The proteomic analysis was from a single first-trimester time point, and longitudinal sampling might reveal dynamic changes in proteomic profiles that further distinguish HDP subtypes. However, we have not performed proteomics on later pregnancy samples at this time. We recognize that there are further ways to categorize the phenotypes of the umbrella clinical diagnoses of HDP, including gestational hypertension versus preeclampsia and preterm (<34 weeks of gestation) HDP. These questions have been interrogated by other studies, which have found partially overlapping molecular signatures between gestational hypertension and preeclampsia.^39–41^ The primary question of interest in this analysis was related to the timing of HDP for all clinical conditions classified as HDP. As such, additional breakdown into gestational hypertension and preeclampsia within each of the timing groups would result in small sample sizes within each group and would significantly limit our power. Finally, the pathway analyses, while biologically informative, are based on curated databases that may not fully capture pregnancy-specific biology. Accordingly, the canonical pathway names (e.g., “SPINK1 Pancreatic Cancer Pathway”, “Airway Pathology in COPD”) reflect the databases in which these pathways were originally characterized rather than their relevance to pregnancy.

## PERSPECTIVES

There are differences in protein expression in early pregnancy based on whether HDP is diagnosed in the antepartum, intrapartum, or postpartum period. In our analysis, the proteomic signature of antepartum HDP aligns with known angiogenic imbalance pathways, while that of intrapartum HDP aligns with altered inflammatory pathways, and that of postpartum HDP aligns more with cardiovascular remodeling and complement-mediated cardiac dysfunction pathways. These findings raise the possibility that there may be distinct mechanistic phenotypes that could uniquely inform diagnostic and therapeutic targets for HDP.

## NOVELTY AND RELEVANCE

1. What is new?

- There are differences in protein expression in early pregnancy based on whether hypertensive disorders of pregnancy (HDP) are diagnosed antepartum, intrapartum, or postpartum.
2. What is relevant?

- Proteomic signature of antepartum HDP aligns with known angiogenic imbalance pathways
- Intrapartum HDP aligns with altered inflammatory pathways
- Postpartum HDP aligns with cardiovascular remodeling and complement-mediated cardiac dysfunction pathways.
3. Clinical/Pathophysiological Implications?

- These findings raise the possibility that there may be distinct mechanistic phenotypes that could uniquely inform diagnostic and therapeutic targets for HDP.

## Data Availability

All data produced in the present study are available upon reasonable request to the authors

